# Improving and Interpreting Surgical Case Duration Prediction with Machine Learning Methodology

**DOI:** 10.1101/2020.06.10.20127910

**Authors:** Jesyin Lai, Ching-Chieh Huang, Shu-Cheng Liu, Jhao-Yu Huang, Der-Yang Cho, Jiaxin Yu

## Abstract

Hospitals have encountered challenges in performing efficient scheduling and good resource management to ensure a high quality of healthcare is provided to their patients. Operating room (OR) scheduling is one of the issues that has gained our attention because it is related to workflow efficiency and critical care of hospitals. Automatic scheduling and high predictive accuracy of surgical case duration have a critical role in improving OR utilization. To estimate surgical case duration, most hospitals might rely on historical averages based on a specific surgeon or a specific procedure type obtained from electronic medical record (EMR) scheduling systems. However, the low predictive accuracy with EMR data leads to negative impacts on patients and hospitals, such as rescheduling of surgeries and cancellation. This study aims to improve and interpret the prediction of surgical case duration with machine learning (ML) methodology. A large data set containing 170,748 surgical cases (from Jan 2017 to Dec 2019) was obtained from a hospital, and it covered a broad variety of details on patients, surgeries, specialties and surgical teams. In addition, a more recent data set with 8,672 cases (from Mar to Apr 2020) was available to be used for time-wise evaluation. Historical averages were computed from the EMR data for surgeon- or procedure-specific cases, and served as baseline models for comparison. Subsequently, models were built with linear regression, random forest and extreme gradient boosting (XGB) algorithms, and were evaluated with R-square (*R*^2^), mean absolute error (*MAE*), percentage overage (actual duration longer than prediction), underage (shorter than prediction) and within (absolute duration differences falling within minimum(maximum(15 %,15 min), 60 min) of prediction). The XGB model was superior to the other models, achieving a higher *R*^2^ (84 %) as well as a lower *MAE* (31.1 min) and inaccurate percentage (24.4 %). In addition, XGB predictions were analyzed with Shapley additive explanations (SHAP). SHAP interpretation on complex cases (e.g. containing more than 2 procedures) unraveled that older primary surgeons took shorter time to complete the surgery and primary surgeons with longer previous surgical time within a week took more time to complete the surgery. Longer durations were utilized when patient’s hypertension status was unknown. Meanwhile, SHAP interpretation on model loss showed that the loss values of elder primary surgeons increased for cases with larger deviations in prediction suggesting additional information related to surgeon is required for model improvement. Overall, this study applied ML techniques in the field of OR scheduling to reduce the medical and financial burden for healthcare management. The results revealed the impact of main factors (e.g. anesthesia, procedure types, no. of procedure) and interaction effects (e.g. no. of procedure x primary surgeon’s age) in surgical case duration prediction as well as identified the feature that contributes to errors in prediction.

## Introduction

It has become increasingly important for clinics and hospitals to manage resources for critical care during the COVID-19 pandemic period. Statistics show that approximately 60 % of patients admitted to the hospital will need to be treated in the operating room (OR) [1], and the average OR cost is up to 2,190 dollars per hour in the United States [2, 3]. Hence, the OR is considered as one of the highest hospital revenue generators and accounts for as much as 42 % of a hospital’s revenue [4, 3]. Based on these statistics, a modern OR schedule and management strategy is not only critical to patients who are in need of elective, urgent and emergent surgeries but is also important for surgical teams to be prepared. Owing to the importance of the OR, improvement of OR efficiency has high priority so that the cost and time spent on the OR is minimized while the utilization of OR is maximized to increase the surgical case number and patient access [5].

In a healthcare system, numerous factors are involved in affecting OR efficiency, for example, patient expectation and satisfaction, interactions between different professional specialties, unpredictability during surgeries, surgical case scheduling, etc. [6]. Although the OR process is complex and involves multiple parties, one way to enhance OR efficiency is by increasing the accuracy of predicted surgical case duration. Over- or underutilization of OR time often leads to undesirable consequences such as idle time, overtime, cancellation or rescheduling of surgeries, which may induce a negative impact on the patient, staff and hospital [7]. In contrast, high efficiency in OR scheduling not only contributes to a better arrangement for the usage of the OR and resources but can also lead to a cost reduction and revenue increase since more surgeries can be performed.

Currently, most hospitals schedule surgical case duration by employing estimations from the surgeon and/or averages of historical case durations, and studies show that both of these methods have limited accuracy [8, 9]. For case lengths estimated by surgeons, factors including patient conditions and anesthetic issues might not be taken into consideration. Moreover, underestimation of case duration often occurs because surgeon estimations are usually made by favoring maximizing block scheduling to account for potential cancellations and cost reduction. Furthermore, operations with higher uncertainty and unexpected findings during surgery add difficulties and challenges to case length estimation [8]. Historical averages of case duration for a specific surgeon or a specific type of surgery obtained from electronic medical record (EMR) scheduling systems have also been used in hospitals. However, these methods have been shown to produce low accuracy due to the large variability and lack of the same combination of factors in the preoperative data available on the case that is being performed [10].

To improve the predictability, researchers have utilized linear statistical models, such as regression, or simulation for surgical duration prediction and evaluation of the importance of input variables [11, 12, 13]. However, a common shortcoming of these studies is that relatively fewer input variables or features were used in their models than in alternative approaches due to the limitation of statistical techniques in handling too many input variables. Recently, machine learning (ML) has been shown to be powerful and effective in aiding health care management. Master et al. (2017) trained multiple ML models, including decision tree regression, random forest regression, gradient boosted regression trees and hybrid combinations, to automate prediction and classification of pediatric surgical durations [14]. Ensemble algorithms, implementing least-squares boosting and bagging models with ML, developed by Shahabikargar et al. (2017) were shown to reduce the error by 55 % compared to the original error [7]. With the use of a boosted regression tree, Zhao et al. (2019) increased the percentage of accurately booked cases for robot-assisted surgery from 35 % to 52 %. Bartek et al. (2019) reported that they were able to improve predicted cases within 10 % of the threshold tolerance from 32 % to 39 % using an extreme gradient boosting model [15]. Nonetheless, these ML studies included only 5-12 different types of procedures and specialties to train their ML models, which may limit the generalization of these models.

In this study, more than 170,000 cases were obtained from China Medical University Hospital (CMUH) containing 743 types of procedures across 25 different specialties. From the original data, we further analyzed the working time of primary surgeons and computed their total number of previous surgeries and the total minutes spent on previous surgeries within 24 hr as well as within the last 7 days. Since surgeons’ working performance might be affected by previous events, surgical cases performed by the same primary surgeon, especially within 24 hr, should not be considered as totally independent and unrelated. Hence, previous surgical counts and working time obtained from surgeons’ data were included as additional features in our ML model training to account for their influences on surgical case duration. We hypothesize that these features impose significant influences on surgical case duration and may aid in improving the performance of a trained ML model.

## Results

### Model development and evaluation

Approximately 17 % of the cases were excluded from the original data from Jan 1, 2017, to Dec 31, 2019, based on the exclusion criteria mentioned in Fig. S2 (Supplementary info). Therefore, 142,445 cases containing more than 743 procedural categories and 25 specialties were included for predictive model development and evaluation. Furthermore, a recent data set collected from Mar 1 to April 30, 2020, (7,231 cases after exclusion) was used in the time-wise evaluation to verify the robustness of the model in making predictions.

The results of all the metrics used to evaluate model performance on training, internal validation and time-wise testing sets are reported in Table S1 of the Supplementary info. Fig. 1 summarizes the results of the model evaluation on the time-wise testing set. The figure shows that the average model for the surgeon-specific scenario was not a good estimate for surgical case duration. The average model for the procedure-specific scenario had a lower percentage underage (actual duration shorter than prediction) and overage (longer than prediction) than the surgeon-specific average model. These differences were due to an extensive procedure classification in the procedure-specific model. However, the percentage underage was still quite high. Since no other information is taken into consideration in both of the average models except the duration of surgical cases that happened in the past, these models usually exhibit prediction bias and low accuracy.

**Figure 1:**
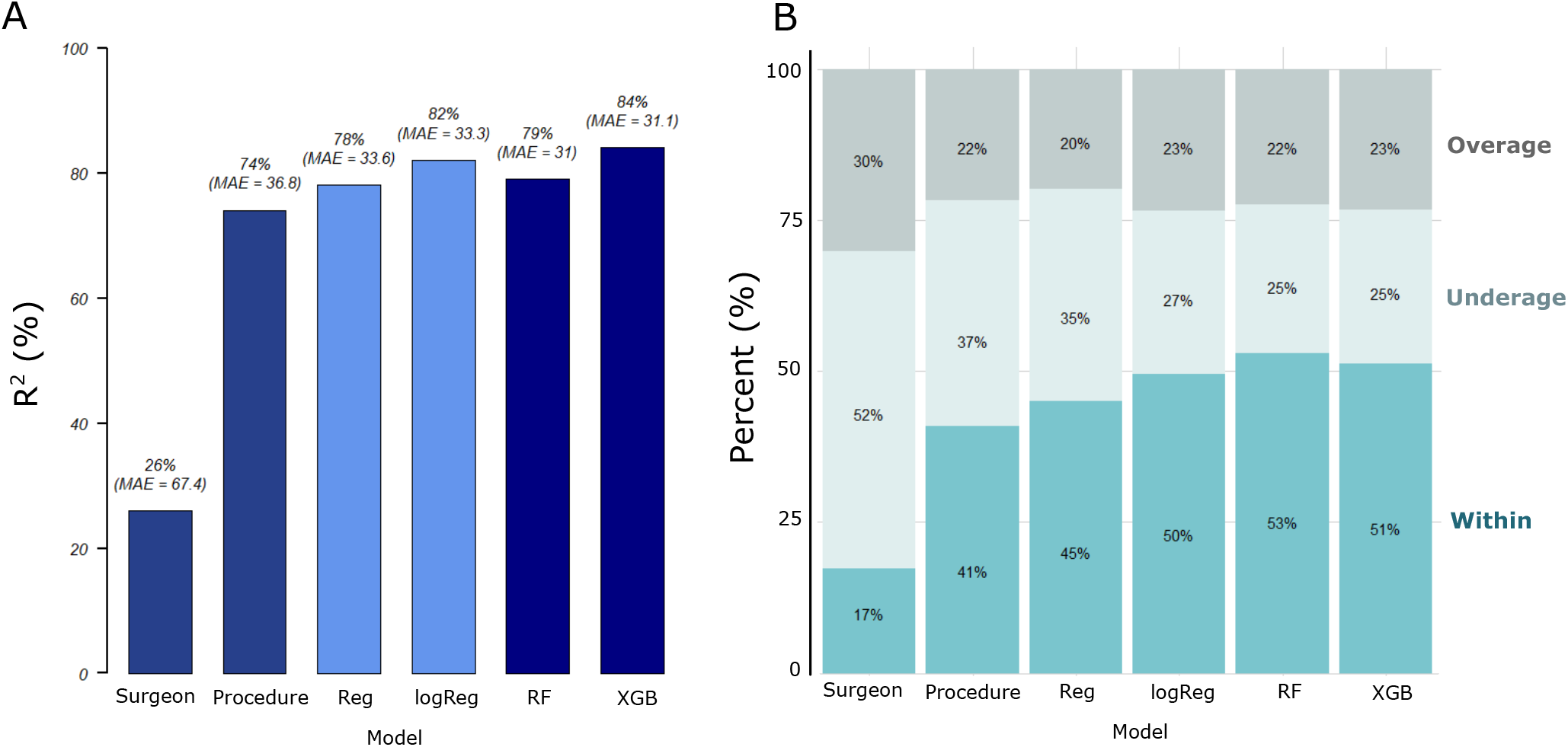
Machine learning algorithms improved the prediction accuracy of surgical case duration based on the results of time-wise model evaluation. The performance of all models was evaluated on a time-wise testing set (data not included in the original data set for ML model training) by using (A) *R*^2^, mean absolute error (*MAE*) as well as (B) percentage overage (actual duration longer than prediction), underage (shorter than prediction) and within (absolute duration differences falling within minimum(maximum(15 %,15 min), 60 min) of prediction).

We first fitted the linear regression (Reg) model by including all the input variables shown in Table 1. The evaluation metrics reported a higher percentage within than the average models on the time-wise testing sets (Fig. 1). There was an increase in the *R*^2^ value, indicating that predictive performance of the model improved when other information was taken into consideration. Since the distribution of the surgical case duration was found to be non-normal (Fig. S1A), we tested Box-Cox transformation on the Reg model. The results showed that the measure of goodness of fit was maximum when the optimal value for transformation was close to 0 (results not shown). Therefore, we applied log-transformation on the surgical case duration. The log-transformation is a special case of the Box-Cox transformation [16]. Moreover, by applying log-transformation, the differences between shorter durations were expanded while the differences between longer durations were reduced. This is because the slope of the logarithmic function is steeper for smaller values but moderate for larger values. When we log-transformed surgical case duration and reran a regression model (i.e., logReg), the performance of the logReg model outperformed the Reg model.

**Table 1:** Preoperative data with 19 predictor variables were used as inputs for model development. The predictor variables can be categorized by relationship to patient, surgical team, operation, facility and surgeon’s prior events. These predictor variables were selected based on the significance (p-value *<* 0.05) of their correlations with the outcome using a regression analysis. The text in the parentheses is the code name of the corresponding feature variable that was used during model development and interpretation. BMI classification was performed following the standard for Asians and the categories are listed in Table S3. ICD: International Classification of Diseases; ASA: American Society of Anesthesiologists; BMI: Body mass index.

| Patient | Surgical team | Operation | Facility | Primary Surgeon's Prior Events |
| --- | --- | --- | --- | --- |
| Age (age) | Surgeon team size (TeamSize) | Procedure type (Proced) | Day of the week (weekday) | No. of previous surgeries performed by the surgeon within 24 hr (Opcount_1d) |
| Gender (SexName) | Specialty (DivNo) | Anesthesia type (AnaValue) | Time of day (TimeofDay) | Total surgical minutes performed by the surgeon within 24 hr (Optoltime_1d) |
| ICD code (Diag) | Primary surgeon's age (Dr_age) | No. of procedure (Nproced) |  | No. of previous surgeries performed by the surgeon within the last 7 days (Opcount_7d) |
| In- /out-patient (OpType) |  |  |  | Total surgical minutes performed by the surgeon within the last 7 days (Optoltime_7d) |
| ASA status (ASA) |  |  |  |  |
| Hypertension (Hypertension) |  |  |  |  |
| BMI category (BMI) |  |  |  |  |

Although the performance of the logReg model was better than the Reg and the average models, an assumption of a linear relationship between the target and input variables was applied in both the Reg and the logReg models. The relationship between the target and input variables is usually nonlinear in a real-world situation. ML algorithms are helpful in making predictions in a more complicated scenario. The random forest (RF) model is the first ML model that we built in this study. There was a slight improvement in *MAE* compared to that of the logReg model. However, overfitting was observed in the RF model as performance of the training set was better than the internal validation and time-wise testing sets. An XGB model was subsequently developed to find out if the predictive performance would be better. Although the results of MAE, underage, within and overage were similar, the XGB model outperformed the RF model in terms of *R*^2^. Hence, the XGB model was chosen as the best model and was used in subsequent analysis.

In addition to the three key metrics, we studied the accumulative inaccuracy of all the models by using the time-wise testing set. The total prediction error (in minutes) and the corresponding inaccurate percentage were calculated (Table 2). The actual total minutes represent the sum of surgical case durations for 7,231 cases in the time-wise testing set. The inaccurate percentage was derived from the percentage of total prediction error divided by the actual total minutes. The outcome shows that the inaccurate percentages of both the ML models (RF and XBG) were the lowest. Since the XGB model was not overfitted and had a higher *R*^2^ value, the XGB model was applied to improve the efficiency of OR scheduling. In addition to looking at the overall performance across all specialties, we compared the performance of the XGB model by further breaking it down to the specialty level on the time-wise testing set. Fig. S3 (Supplementary Info) shows the number of cases that were predicted as overage, underage and within for each specialty in the time-wise testing set. Most specialties had more cases that were predicted as within than as overage and underage. There were some exceptional specialties which had fewer cases of within because the total case numbers in these specialties were low. Moreover, the mean case durations were long for bariatric and metabolic as well as pediatric dentistry. As these cases were rare and required longer duration, this rendered difficulties for the model to predict accurately.

**Table 2:** The extreme gradient boosting (XGB) and random forest (RF) model produced the lowest percentage of cumulative inaccuracy among all the other models. Cumulative differences between actual and predicted case durations for all the models are shown in this table.

|  | Actual | Surgeon-specific | Procedure-specific | Reg | logReg | RF | XGB |
| --- | --- | --- | --- | --- | --- | --- | --- |
| **Total minutes** | 920,374 | 910,190 | 938,154 | 945,001 | 903,983 | 875,144 | 892,707 |
| **Total prediction error in minutes** |  | 487,357 | 266,185 | 243,233 | 240,980 | 224,476 | 224,643 |
| **Inaccurate percentage (%)** |  | 53 | 28.9 | 26.4 | 26.2 | 24.4 | 24.4 |

Subsequently, we plotted Bland-Altman (BA) plots using the time-wise testing set for the average models (surgeon- and procedure-specific) and the XGB model (Fig. 2). The BA plots for these three models were plotted on y-axes in two different scales, one is (actual-prediction) (Fig. 2A-C) and the other is (actual-prediction)/prediction (Fig. 2D-F). Histograms for x- and y-axes were also plotted alongside the corresponding BA plots in Fig. 2A-C. The BA plots and the histograms for y-axes clearly show that predictions of the XGB model, especially the shorter durations, were closer to the actual durations compared to predictions of the two average models. When duration differences were divided by predictions (Fig. 2D-F), they represent the deviations of differences from the predictions. This helps quantifying to what extent the differences occurred relative to predictions. In contrast to the average models, BA plot of the XGB model (Fig 2F) reveals deviations were much smaller and less scattered, including cases that were considered as overage and underage as well as long-duration cases. As a result, this demonstrates that the XGB model is more accurate than the average models in predicting surgical case duration.

**Figure 2:**
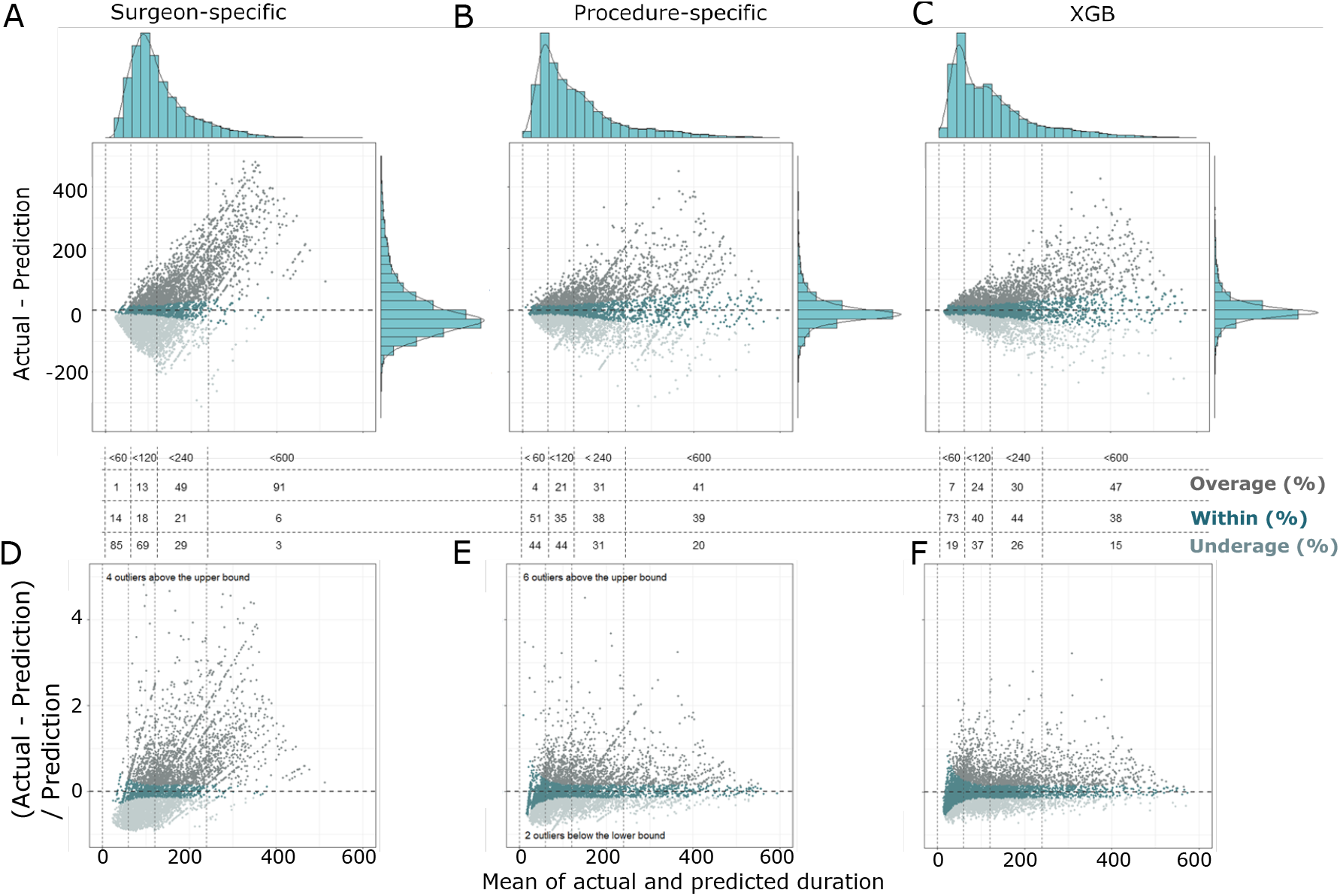
Predictions produced by the extreme gradient boosting (XGB) model, especially the shorter durations, were closer to the actual surgical case durations compared to predictions produced by average models. The Bland-Altman plots (with histograms for x- and y-axes) for average models of (A) surgeon- and (B) procedure-specific scenarios, and (C) the XGB model revealed that differences between actual and predicted durations in the XGB model were smaller. Bland-Altman plots with differences versus predicted durations for (D) surgeon- and (E) procedure-specific scenarios, as well as for (F) the XGB model illustrated that actual and predicted duration differences of the XGB model were smaller in deviations from the predictions. Percentage overage, within and underage for cases falling within 0-60 min, 60-120 min, 120-240 min and 240-600 min of these three models, respectively, were reported in the figure. Data points were colored according to the classification of overage, within and underage.

### Model interpretation

To uncover the global importance and the impact of each feature on the XGB model output, we extracted weighted feature gain (WFG) and applied Shapley additive explanations (SHAP) to explain the model. WFG was computed based on the reduction in model accuracy when the variable was removed. This value serves as an indication of how important the variable is in improving the purity of a decision tree branch [17, 18]. A higher WFG percentage indicates that the variable is more important. Although WFG is widely used as the basis for feature selection methods, it may not be consistent with other feature attribution methods, such as split count or permutation [19]. In contrast, SHAP adopts the classical Shapley values estimation methods, which satisfy the desirable properties of local accuracy, missingness and consistency [20, 19]. It explains model output by computing the contribution of each feature to the prediction. Fig. 3A reports feature importance based on WFG while Fig. 3B shows feature impact on model output based on SHAP value. Both measures reveal that anesthesia type, procedure type, no. of procedure, hypertension and specialty were the 5 most important features in the XGB model. Notably, 3 of the top 5 important variables were attributed to operative information (i.e., anesthesia type, procedure type and no. of procedure). Moreover, two of the features that we computed from the surgeon data (i.e., total surgical minutes and the number of previous surgeries performed by the surgeon within the last 7 days) were included within the top 10 list of both measures. Although there were some slight differences in the ranking order of feature importance, SHAP value is more informative as it provides quantification and visualization of the impact (negative or positive) of each feature on the final output for each case as well as the variations in feature contribution relative to changes in feature value (Fig. 4A).

**Figure 3:**
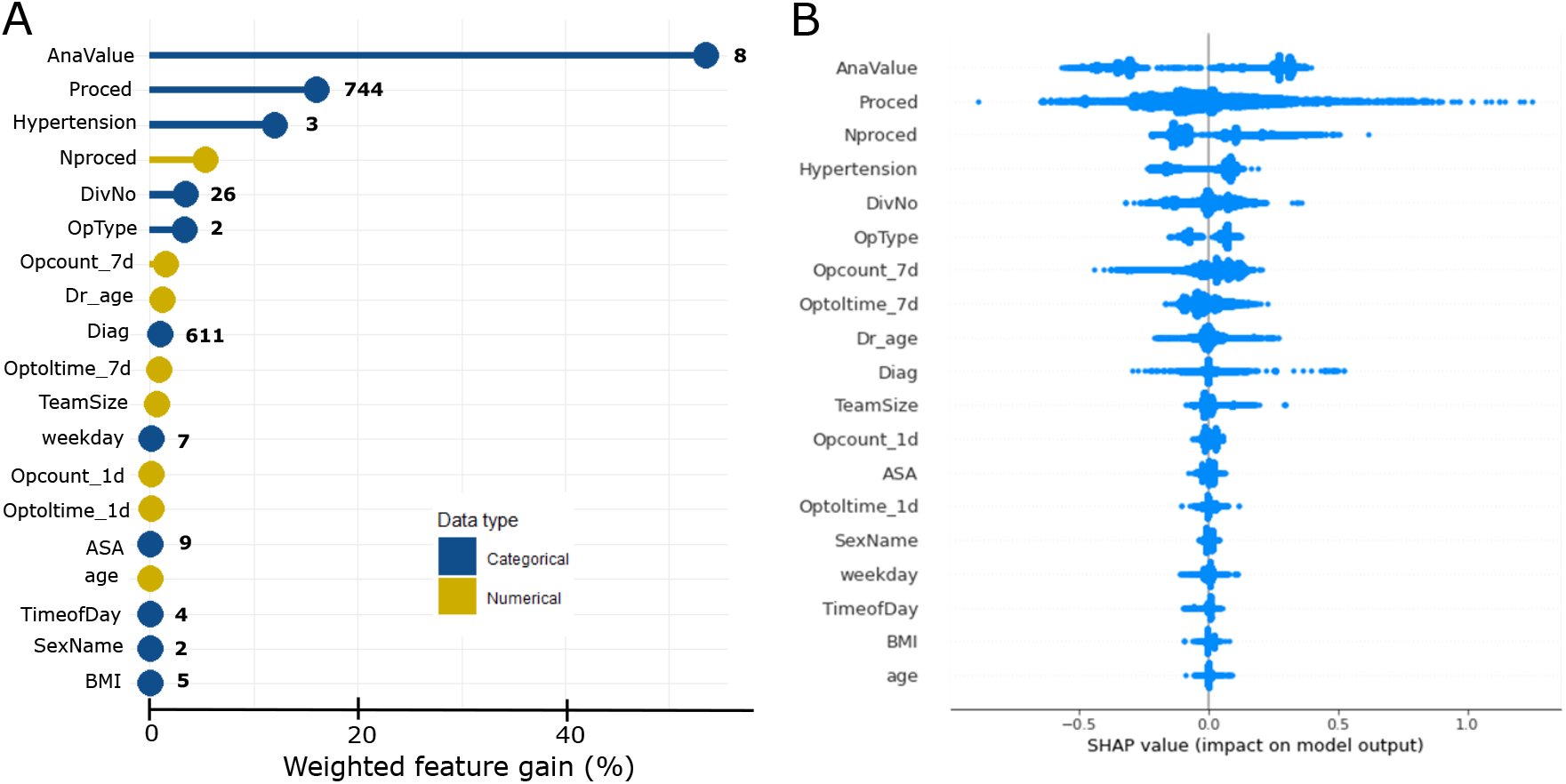
Anesthesia type, procedure type, number of procedure, hypertension and specialty were the top 5 important features used by the extreme gradient boosting (XGB) model to make predictions. Features were arranged and ranked in descending order according to (A) weighted feature gain or (B) SHAP value. In panel (A), categorical features are represented by blue markers, while numerical features are represented by yellow markers. The numbers of categories for categorical features are also shown beside the markers. The results of SHAP value show that total surgical minutes and number of previous surgeries performed by the surgeon within the last 7 days (‘Optoltime_7d’ and ‘Opcount_7d’) had important contributions to model output. Specific code names are used to represent the full names of features in the figure. The features’ full names can be referred to Table 1.

**Figure 4:**
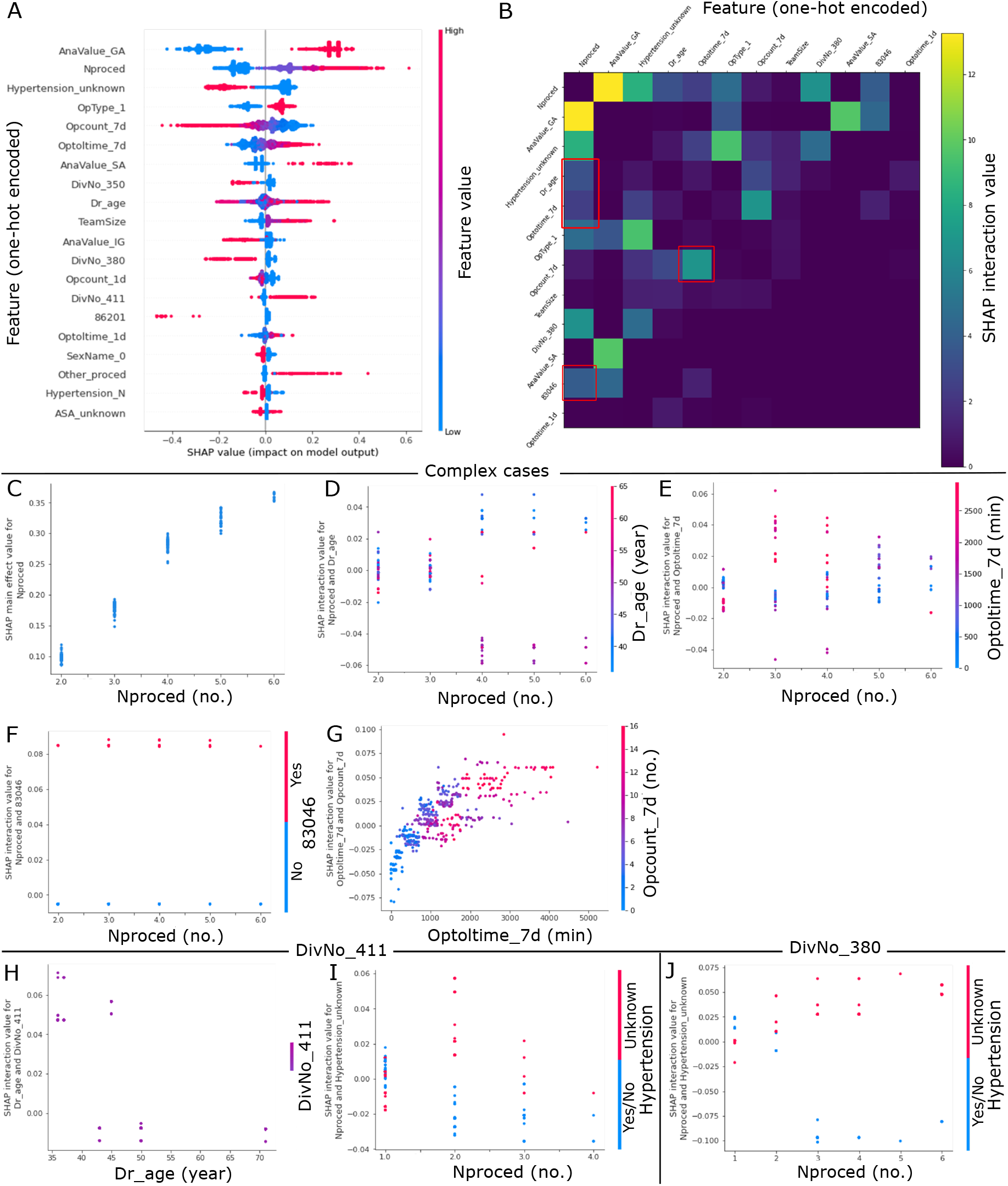
Shapley additive explanations (SHAP) interaction value unravels interactions between various one-hot encoded features. (A) A summary plot of 20 one-hot encoded features with the largest average SHAP value magnitude for the extreme gradient boosting (XGB) model. Each dot corresponds to a case in the time-wise testing set. No. of procedure (‘Nproced’), anesthesia type of spinal anesthesia (‘AnaValue SA’), Team size (‘TeamSize’), specialty of plastic and reconstruction (‘DivNo 411’), and other procedure (‘Other proced’) had long positive tails. These indicate that these features tended to increase model output (longer surgical case duration) when feature values were either high (for numerical feature) or 1 (for categorical feature). (B) A heatmap summarizing SHAP interaction values of one-hot encoded features for complex cases extracted from the testing set. One-hot encoded features with top 12 largest SHAP interaction values are shown. Note that the interaction effects are shown off-diagonal and symmetrically. Higher and interesting interaction effects are highlighted with red boxes. (C) SHAP main effect value for no. procedure (‘Nproced’). Larger no. of procedure had increased impact on model output. (D) SHAP interaction value for no. of procedure and primary surgeon’s age (‘Dr_age’). For cases with 4-6 procedures, older primary surgeons were associated with shorter case durations. (E) SHAP interaction value for no. of procedure and total surgical minutes of previous surgeries performed by the surgeon within the last 7 days (‘Optoltime_7d’). For cases with 3-5 procedures, primary surgeons with larger ‘Optoltime_7d’ were associated with longer case durations. (F) SHAP interaction value for no. of procedure and procedure type of spinal infusion surgery with spinal instrumentation equal or less than 6 segments (‘83046’). Regardless of the no. of procedure, longer durations were consumed for cases with procedure code of ‘83046’. (G) SHAP interaction value for total surgical minutes (‘Optoltime_7d’) and number (‘Opcount_7d’) of previous surgeries performed by the surgeon within the last 7 days. The impact of ‘Optiltime_7d’ on case durations was affected by changes in ‘Opcount 7d’. (H) and (I) were plotted for surgical cases belonged to the specialty of plastic and reconstruction (‘DivNo_411’ = 1), while (J) was plotted for surgical cases belonged to the ophthalmology specialty (‘DivNo_380’ = 1). (H) SHAP interaction value for primary surgeon’s age and cases belong to the specialty of plastic and reconstruction. Younger primary surgeons took longer time to complete the surgery. (I) and (J) SHAP interaction value for no. of procedure and hypertension status. In both specialties of plastic and reconstruction, and ophthalmology, cases with hypertension status of unknown were associated with longer durations as no. of procedure increased.

While SHAP summary plot provides global interpretability reflecting the general behavior of the features in the model, we also investigated the specific behavior of the features in a subset of model predictions. As complex cases with more procedure numbers and larger team size may benefit more from this ML predictive approach, we extracted complex cases from the time-wise testing set for local interpretability. From the SHAP summary plot of one-hot encoded features in Fig. 4A, we observed that cases with anesthesia type of general anesthesia (GA) and out-patient cases were associated with higher model output (longer duration) in addition to larger no. of procedure and surgeon team size. In contrast, cases belong to the specialties of otolaryngology, head, and neck (‘DivNo 350’), and ophthalmology (‘DivNo 380’) were associated with lower model output (shorter duration). Therefore, we selected complex cases by setting the criteria to be no. of procedure *≥* 2, team size *≥* 2, anesthesia type = GA, and not in the specialties of otolaryngology, head, and neck, and ophthalmology. Fig. 4B is a heatmap of SHAP interaction values of one-hot encoded features for the extracted complex cases. Only one-hot encoded features with top 12 largest SHAP interaction values are reported in the figure. Features with higher and interesting interactive effects were highlighted in red boxes and their SHAP interaction values were plotted in Fig. 4D-G. No. of procedure had the highest total SHAP interaction value among all other features. The plot of its SHAP main effect value (Fig. 4C) unravels that an increase in no. of procedure was highly associated with an increase in predicted duration. Features, including primary surgeon’s age (Fig. 4D), total surgical minutes performed by the surgeon within the last 7 days (Fig. 4E), and procedure type of spinal infusion surgery with spinal instrumentation equal or less than 6 segments (‘83046’, Fig. 4F), had higher SHAP interaction values with no. of procedure. From these SHAP interactive value plots, a few interesting phenomena were observed. For complex cases with 4-6 procedures, older primary surgeons were associated with shorter surgical case durations. On the other hand, primary surgeons with longer previous surgical minutes within the last 7 days were associated with longer durations for complex cases with 3-5 procedures. For procedure type of spinal infusion surgery with code ‘83046’, surgical case durations were longer regardless of the no. of procedure. Total surgical minutes (‘Optoltime_7d’) also interacted strongly with total no. (‘Optolcount_7d’) of previous surgeries performed by the surgeon within the last 7 days (Fig. 4G). This reflects that primary surgeons with higher ‘Optoltime_7d’ and higher ‘Optolcount_7d’, but not moderate ‘Optoltime_7d’ and higher ‘Optolcount_7d’, were associated with longer durations. In summary, these interactive effects suggest that older primary surgeons took shorter time to finish complex cases and primary surgeons with longer previous surgical minutes tended to take longer time to complete complex surgeries. Moreover, complex cases with procedure code ‘83046’ consumed longer durations.

When we looked at SHAP interaction value for cases belong to the specialty of plastic and reconstruction (‘DivNo_411’), similar effect of primary surgeon’s age was also observed. Fig. 4H shows that primary surgeons with younger age had larger positive impact on surgical case duration than those with older age implying that younger primary surgeons took longer time to perform the surgery. Moreover, for cases extracted from this specialty and the specialty of ophthalmology (‘DivNo_380’), cases with more no. of procedure and patient’s hypertension status of unknown were associated with longer durations. However, this was not observed in cases with one procedure. These suggest that hypertension might be an important factor affecting surgical case duration. Hence, hypertension status of unknown might added in uncertainties and increased duration of cases with two or more procedure numbers.

In addition to global and local interpretability on model prediction, it is also important to find out factors that contribute to errors in model prediction. This may subsequently help to debug and improve the model. SHAP loss value is a tool in SHAP package that can be used to find out each feature contribution to the model loss. Loss can be seen as a magnitude between the actual values and the predicted values. A larger loss implies a larger error in model output. Hence, lower model loss is preferable as this indicates that the predicted values are closer to the actual values. As Fig. 2C reveals that cases with shorter durations had less errors than longer durations in the XGB model, we extracted cases with predicted durations falling in the range of 180-600 min from the testing set and investigated which features contribute to higher errors, i.e. higher loss. Subsequently, we selected cases with absolute duration differences less than 30 min (clinically not significant differences) or more than 90 min (large differences). Fig. 5A and B are summary plots of the top 10 one-hot encoded features with the highest average SHAP loss value magnitude for cases with small and large differences, respectively. The SHAP loss value of primary surgeon’s age (‘Dr_age’) was noticed because this feature had a longer positive tail, i.e. higher contribution to the model loss, in cases with large differences than cases with small differences. Meanwhile, other features had long negative tails implying that these features contributed to the reduction in model loss. We then focused on cases with predicted durations of 180-600 min to look at the distribution of SHAP loss value for one feature. We sorted all the cases according to their absolute differences normalized to predictions (which quantify the deviations of differences from the predictions) and plotted Fig. 5C, which manifests deviations (y-axis) were larger for cases with larger index numbers (x-axis). The SHAP loss value for the feature of primary surgeon’s age in Fig. 5D reveals a significant inconsistency (grey vertical line) in loss value as case index increases (i.e., cases with larger deviations). The grey vertical dashed line indicates the location where the t-test (with Bonferroni correction) has a p-value less than 0.05/(no. of comparisons). The t-test was automatically and repeated conducted to compare two data samples splitting with an increment of 50 data points when plotting the figure with a function in the SHAP python package. When we manually compared SHAP loss values of the first 400 and last 400 cases with primary surgeon’s age older than 65 years (Fig. 5D) using a one-sided t-test, the SHAP loss values of the latter were significantly (t = −2.64, p-value = 0.01) higher. These results suggest that primary surgeon’s age is one of the factors contributing to the increase in prediction error. In contrast, the SHAP loss value for the feature of no. of procedure in Fig. 5E were consistent for cases with smaller and larger deviations. Although the mean SHAP loss value magnitude of no. of procedure was the largest, its contribution on model loss was observed to be not associated with the increase in error made by the model.

**Figure 5:**
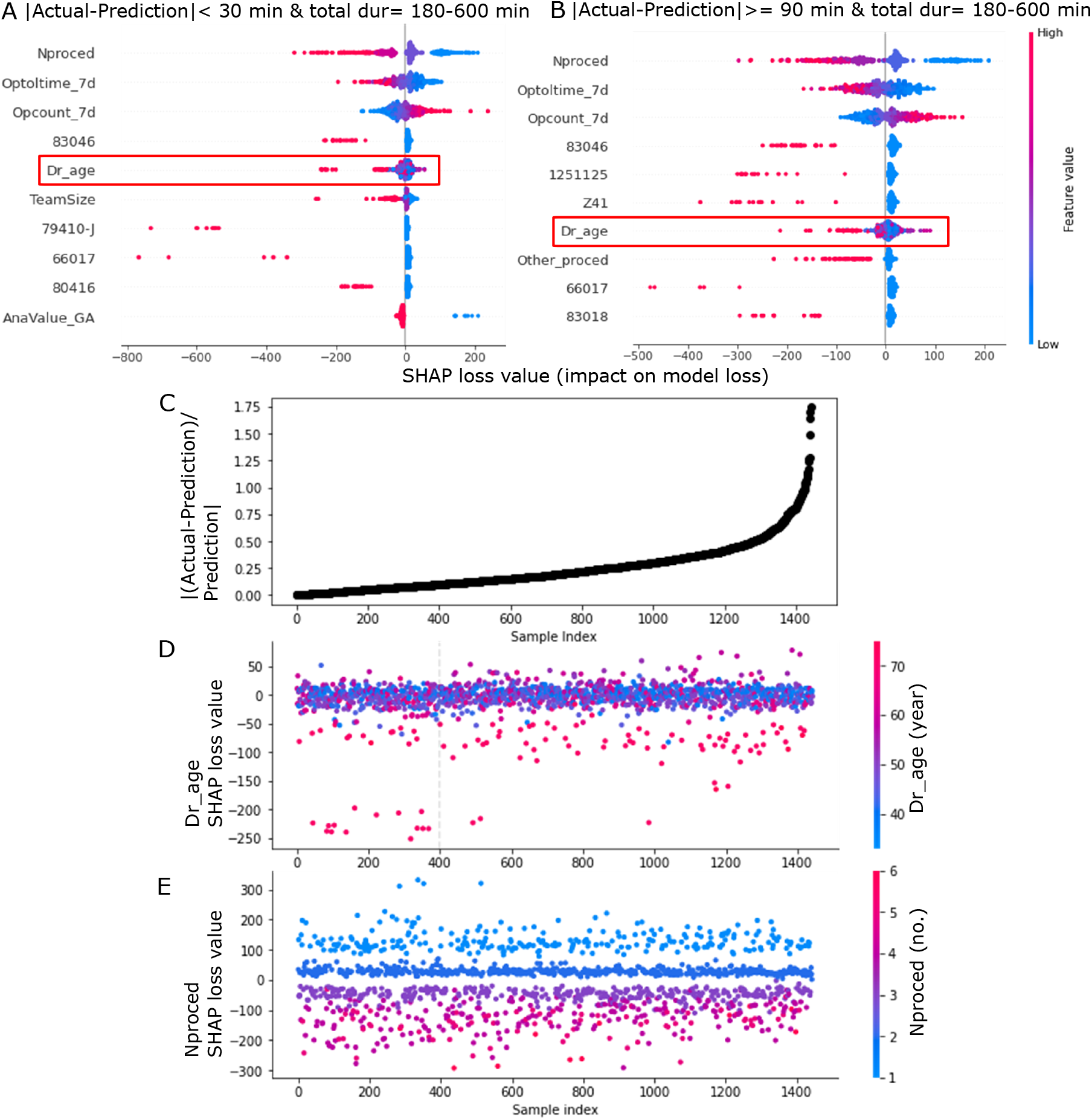
Shapley additive explanations (SHAP) loss value aids in identifying features with larger contribution to the model loss. Cases with predicted durations of 180-600 min were extracted from the time-wise testing set. Subsequently, cases with absolute duration differences (| actual - prediction|) less than 30 min or more than 90 min were selected. Summary plots of 10 one-hot encoded features with the largest average SHAP loss value magnitude for (A) cases with differences less than 30 min (clinically not significant differences) and (B) cases with differences more than 90 min (large differences). Each dot corresponds to a case. The SHAP loss values of primary surgeon’s age (‘Dr_age’) are highlighted in red boxes in (A) and (B). This feature had a longer positive tail in cases with large differences compared to cases with small differences. (C) All cases with predicted durations of 180-600 min were sorted according to their absolute differences normalized to predictions (| (actual - prediction)/prediction |). The dot plot shows that deviations (y-axis) were larger for cases with larger index numbers (x-axis). (D) The SHAP loss value for the feature of primary surgeon’s age. A grey vertical dashed line indicates the location where the t-test (with Bonferroni correction) has a value less than 0.05/(no. of comparisons), i.e. this reflects the inconsistency of SHAP loss value for the feature. The t-test was automatically and repeated conducted to compare two data samples splitting with an increment of 50 data points when plotting the figure with a function in the SHAP python package. This indicates that primary surgeon’s age is one of the factors contributing to the increase in prediction error. (E) The SHAP loss value for the feature of no. of procedure (‘Nproced’). In contrast to primary surgeon’s age, SHAP loss values for no. of procedure exhibited consistency for cases with smaller and larger deviations. Although the mean SHAP loss value magnitude of no. of procedure was the largest, it was not related to changes in error produced by the model.

## Discussion

Clinical unmet needs related to OR scheduling can be identified based on the aspects of hospital management, surgical supporting staffs, patients and surgeons [21]. From the hospital management’s point of view, it is difficult for them to strike a balance between decreasing idle time and avoiding overtime of staff to maximize OR utility as well as to reduce costs. Idle time is harmful to maintaining a cost effective OR because time available for a surgical procedure to be performed is not being used. Meanwhile, OR over-utilization is 2.5 times more costly than under-utilization [22]. For surgical supporting staffs, they have to work unexpected overtime and under stressful circumstances when actual durations are significantly longer than scheduled surgical case durations. This may lead to job dissatisfaction and higher turnover rate, which may subsequently affect the quality of care and safety provided by supporting staffs as they are overload or overworked. From the perspective of patients, they often do not know when is the start time of their surgeries if the surgeries are not scheduled as the first case in the morning. Hence, prolonged wait times and increased uncertainties may lead to significant patient dissatisfaction. As for surgeons, new surgeons, who are not assigned to have the first case, are more mindful of scheduling accuracy. This is because they may need to wait on a prior case to be finished by a different surgeon when actual durations are longer than scheduled durations. Continued delays in the same OR may also result in lack of staff to cover late cases.

Owing to the above mentioned needs, accurate prediction of surgical case duration plays a vital role in increasing OR efficiency, reducing costs, maintaining hospital reputation, as well as improving patient and surgeon satisfaction. This study not only helps to improve the accuracy of OR case prediction but also provides meaningful insights on how predictions were made by the developed ML model. It has both clinical and technical novelties in the following aspects. For the clinical aspect, we are the first who modeled OR events as dependent events instead of independent. We extracted some additional information from surgeon data, e.g., previous working time and no. of previous surgeries of the primary surgeons within the last 7 days and 24 hr, and this information was taken into consideration during model building. Interpretation on model output unraveled that previous working time and no. of previous surgeries interacted with each other and produced differential effects on surgical case durations. For primary surgeons who performed surgeries containing more than 2 procedures, having longer cumulative previous surgeries time tended to affect their performance on the surgery (Fig. 4E G). Furthermore, while other past studies only reported how they developed and evaluated their predictive models [15, 23, 14, 24, 7], we went further to perform global and local interpretability on model prediction as well as identify factors contributing to errors in model prediction. To this end, older primary surgeons were observed to take shorter time to complete complex surgeries with 4-6 procedures and longer duration was required when patient’s hypertension status was unknown. The phenomenon regarding patient’s hypertension status was identified in more than one specialties suggesting knowing patient’s hypertension status may be critical to surgical time management. Even though older primary surgeon might have more experiences and tended to finish surgeries faster (Fig. 6D H), loss values of primary surgeon’s age older than 65 years were inconsistent compared to younger primary surgeons (Fig. 5D). Therefore, to improve model prediction, we suggest including more detailed information on years of experience for primary surgeons and the role of primary surgeons (e.g. executor or consultant) to be used as features for future ML model development.

For the technical aspect, the data set used in this study contained more than 140,000 cases (after exclusion) and more than 700 different types of surgical procedures, establishing a new benchmark for a massive quantity of data with high diversity. The maximal number of cases that had been used in other studies was in the range of 40,000 to 60,000 [15, 7]. By using a large data set with huge diversity and variability, a more powerful ML model was trained and built. Interpretation on the ML model built with such a large and diverse data set subsequently aids in providing various clinically related information, which may not be disclosed by a simple ML model and is not discussed in the past studies. Moreover, the developed XGB model in this study will be deployed as a stand-alone artificial intelligence (AI) server connected by the EMR of the hospital. When a user schedules a surgery using the EMR system, the EMR delivers input parameters to the AI server, where a prediction of surgical case duration is made, and then the AI server sends the output back to the EMR as a suggestion for the user (refer to Fig. S4 in Supplementary Info).

Currently, surgical cases at CMUH are scheduled according to estimates made by primary surgeons. However, surgeon estimates rely heavily on prior experiences of the surgeons, and many factors beyond expectation will not be taken into consideration. Since there is no formal record on surgeon estimates, we used averages that were calculated based on a specific surgeon or procedure type on the time-wise testing set as our baseline models. The performance of these two average models, as reported in Fig. 1 and Table S1 (Supplementary info), clearly showed that they were poor in predicting surgical case duration. These models also tended to underpredict surgical case duration according to their BA plots in Figs. 2. When 19 feature variables (Table 1) were included in our model development, the *R*^2^, MAE, and percentages of underage, overage and within improved substantially compared to the baseline models. When determining the tolerance threshold for percentage within, we set the criterion to be absolute duration differences falling within minimum(maximum(15 %,15 min), 60 min) of prediction. We applied 15 minutes because *±* 15 minutes is an acceptable periodic range at CMUH to be considered as accurate booking. To avoid having an excessively stringent standard, we applied 15 % because a 15 % error in prediction can typically be adapted by the operational management [14]. For cases that took longer than 400 minutes, 15 % of predicted duration could be more than 1 hour and so we adopted 60 minutes to ensure the threshold was not too lax.

By using regression and ML approaches, we were able to decrease the total prediction error (Table 2) of surgical case duration at CMUH. Among all the models, the performance of the XGB model was considered to be the best because it produced the highest *R*^2^ value and had the lowest inaccuracy. Moreover, the XGB model reduced the total prediction error (in minutes) to 224,643 minutes. Since most ORs usually have multiple cases scheduled per day, the total prediction error represents the cumulative effect of total OR cases in the 2-month period of Mar to April 2020. This cumulative effect may eventually reflect a significant financial advantage in scheduling an additional operation case [25]. This approach would also lead to a significant cost reduction and increase in revenue because ORs are utilized appropriately and efficiently. When we evaluated the case numbers of overage, underage and within predicted by the XGB model at the specialty level, there were more case numbers falling within the acceptable thresholds for most of the specialties in the time-wise testing set (Fig. 1). This finding justifies that the performance of the XGB model can be generalized across specialties.

It has been reported in the past studies that primary surgeons contributed the largest variability in surgical case duration prediction compared to other factors attributed to patients [15, 23, 14]. These studies provide evidence and rationale that more factors relating to primary surgeons should be added as input variables in the training of ML models. Moreover, extensive feature engineering usually improves the quality of ML models and can be independent of the modeling technique itself. As a result, in addition to the primary surgeon’s age, we computed previous working time and number of previous surgeries performed by the same primary surgeons within the last 7 days and 24 hr. These variables extracted from primary surgeon data were significantly (p *<* 0.05) correlated with surgical case duration (see Table S2 in the Supplementary info). The correlation coefficients and the impact distribution (Fig. 6A) of these variables also revealed that a surgical case duration of a primary surgeon may decrease as he or she becomes more familiar with the surgical procedure but may increase if his or her total surgical minutes are too long. Although performing a surgery multiple times on different patients may help a primary surgeon to be more efficient in his or her next operation, a long working time may also lead to lethargy and may affect the primary surgeon’s performance. This explanation overlaps with our observations in Fig. 4E and F in which primary surgeons with longer previous surgical durations tended to take longer time to complete the surgery, especially for complex cases containing 3-5 procedures.

In the methodology of data processing, for predictor variables that contained many categories, we grouped categories that had less than 20 cases into a category named ‘Others’. In addition to reducing the data dimensionality for categorical features, this grouping may aid in the generalization of our model, which implies that our model will still be able to predict the case duration even for operations that are rare. Moreover, our model may be applied to new surgeons and other hospitals since surgeon’s identifier and room number were not included in feature inputs. However, there is still a need to update our model after a while or fine-tune the model to better fit the settings of other hospitals. In terms of timing, we recommend updating the model annually by using surgical cases performed in the most recent 3 years as training data.

One limitation in this study is that we selected predictor variables that could only be extracted from preoperative data. Our ML model still needs to be improved in order to be able to predict surgical case duration dynamically. For example, blood loss during surgery may affect case duration since an unexpected increase in blood loss may cause surgeons to take a longer time to complete the surgery. Therefore, it would be better if intra-operative data are incorporated during ML model development, and the prediction made by the ML model can be updated during surgery. Meanwhile, one main reason that we only selected features which can be obtained pre-operatively is because the goal of building a predictive model is to improve and automate surgery scheduling before surgeries. The model, however, does not serve to affect or restrict surgeons on how much time they would need to complete the surgery. Furthermore, one common issue in all ML studies in terms of predicting surgical case duration, including our study, is that ML models were developed using data from a single site. These ML models have limitations in generalization since the surgical team, facilities and patient populations are different across entities. A custom-made model has to be built for a given organization using training data containing its patients, procedures, surgeons, medical staff, and the facility itself. As a result, the exact same ML model may not perform well when applied to another organization or hospital. The other interesting issue of applying ML or AI in surgical duration estimation is that medical technologies quickly evolve. Hence, how frequently an ML or AI model need to be updated still remains to be answered.

## Conclusion

The XGB model was superior in predictive performance when compared to the average, Reg and logReg models. The total inaccurate percentage of the XGB model was the lowest among the other models. When compared to other ML studies, the XGB model built in this study also had a higher coefficient of determination (*R*^2^) and lower percentages of under- and overprediction [15, 24, 7]. Moreover, this model improves the current OR scheduling method at CMUH, which is based on estimates made by surgeons. We validated the model types using a time-wise testing set in addition to the internal validation set split from the original data used in model training. The results provide the evidence that the model generalized well to the time-wise testing data set even during the COVID-19 pandemic period. When external evaluation is not feasible, time-wise evaluation serves as a useful tool to better validate the predictive power of ML models. In addition to model development and evaluation, global and local interpretation on model output as well as identification of the source of errors in model prediction were conducted in this study. Based on the results, older primary surgeons took shorter time to complete the surgery but primary surgeons with longer previous surgical duration within the last 7 days took more time to complete the surgery. Longer duration was consumed when patient’s hypertension status was unknown. Meanwhile, inconsistency in the loss value of primary surgeons aged above 65 years was found to be associated with increased deviations in case duration. Hence, we propose including additional information relevant to surgeon or surgical team composition, such as years of experience and the role of primary surgeon, to be used as predictor variables for future ML model training.

## Methods

### Data sources

Data for this study were collected retrospectively from the EMR scheduling system of CMUH located in Taichung, Taiwan. The data set covered a broad variety of details about patients, surgeries, specialties and surgical teams. A total of 170,748 cases performed between Jan 1, 2017, and Dec 31, 2019, were used for model development. Additionally, 8,672 cases performed between Mar 1 and April 30, 2020, were used as data for time-wise model evaluation in this study. The proportion based on patient characteristics in the overall data set and time-wise testing set was reported in Table S3 in the Supplementary Info. Over 700 different types of procedures across 25 surgical specialties were included in the training data set. Institutional review board approval (CMUH109-REC1-091) was obtained from CMUH before carrying out this study.

### Exclusion criteria, data processing and feature selection

Emergent and urgent surgical cases were removed since these two types of surgeries cannot be scheduled until they happen. Surgical case duration more than 10 hours or less than 10 minutes were also removed. Surgical records with missing values were excluded. Patients who were pregnant as well as duplicate cases were removed. The exclusion criteria are shown in Fig. S2. This approach resulted in a data set of 142,445 cases that were used for model training and validation. The same criteria were also applied to the data of Mar 1 to April 30, 2020, and 7,231 cases remained after exclusion.

Features were selected from available data sources based on literature review and discussion with surgeons and administrators of CMUH. Although the model performance could be enhanced by some postoperative information (e.g., total blood loss), these parameters cannot be used as features for model training because they were either missing or simply estimated by surgeons before surgery. Therefore, only variables that were available before surgery were selected for model development. Furthermore, the correlations of feature variables with the surgical case duration were checked by performing a regression analysis. Only those variables with significant (p-value *<* 0.05) correlation coefficients were selected as predictor variables for model training.

When visualizing all the categories of procedure type and the International Classification of Diseases (ICD) code, there were hundreds to thousands of categories in these two variables. To reduce the problem of having too many dimensions during one-hot encoding of categorical features, we combined categories that had less than 20 cases in the training set into a category and named it as ‘Others’. Similarly, we combined categories for specialty that had less than 20 cases into the category of ‘Others’.

In addition, since surgical case duration can be related to the performance of surgeons and surgeons’ performance is affected by their working time, we analyzed primary surgeons’ previous surgical events. The number of previous surgeries and total surgical minutes performed by the same primary surgeons within the last 7 days and 24 hr. Together, 19 predictor variables were included for predictive model building in this study. These predictors can be categorized into 5 groups: patient, surgical team, operation, facility and primary surgeon’s prior events (see Table 1).

### Model development and training

We applied multiple ML methods for surgical case duration prediction. Surgical case duration (in minutes) is the total period starting from the time the patient enters the OR to the time of exiting the OR. Historical averages of case durations based on surgeon-specific or procedure-specific data from EMR systems were used as baseline models for comparison in case duration prediction. At the beginning, we performed multivariate linear regression (Reg) to predict surgical case duration. However, when we evaluated the distribution of surgical case duration, it was observed to be skewed to the right (Fig. S1 in the Supplementary info). We performed a logarithmic transformation on the surgical case duration to reduce the skewness. The model built from log-transformed multivariate linear regression (logReg) outperformed Reg in all evaluation indexes. Subsequent ML algorithms were also trained by using the log-transformed case duration as the target.

The first ML algorithm that we tested was random forest (RF), a tree-based supervised learning algorithm. RF uses bootstrap aggregation or a bagging technique for regression by constructing a multitude of decision trees based on training data and outputting the mean predicted value from the individual trees [26]. Tree-based techniques were suitable for our data since they include a large number of categorical variables, e.g., ICD code and procedure type, of which most were sparse. The extreme gradient boosting (XGB) algorithm is the other supervised ML algorithm that was tested for comparison to RF. Recently, the XGB algorithm has gained popularity within the data science community due to its ability in overcoming the curse of dimensionality as well as capturing the interaction of variables [27].

XGB is also a decision tree-based algorithm similar to the RF algorithm. However, the XGB and RF algorithms are different in the way that the trees are built. It has been shown that XGB performs better than RF if parameters are tuned carefully; otherwise, it would be more likely to overfit if the data are noisy [28, 29]. For both RF and XGB algorithms, we adopted a 5-fold cross-validation strategy to tune the best hyperparameters, e.g. no. of estimators, maximum of depths, etc. For the XGB model, the best hyperparameters were number of estimators = 535, *η* = 0.5 (step size shrinkage to prevent overfitting), maximum of depths = 3, *γ* = 0.3 (minimum loss reduction, where a larger *γ* represents a more conservative algorithm) and *α* = 1 (L1 regularization weighting term, where a larger value indicates a more conservative model). For the RF model, the best hyperparameters were number of estimators = 150, minimum samples split = 10, minimum samples leaf = 2 and maximum depth = 125.

A data-splitting strategy was used in the training for all the models. We randomly separated the data into training and validation subsets at a ratio of 4:1. The training data were used to build different predictive models as well as to extract important predictor variables. The validation data were used for internal evaluation of the models. In addition to interval evaluation, time-wise evaluation on all the models was performed using data from Mar 1 to Apr 30, 2020. These data were not included in the original data set for ML model training. The results obtained from time-wise evaluation are better in verifying the robustness of the trained model in making an accurate prediction since the data are temporally segregated from the original data. Historical averages of case duration for surgeon- or procedure-specific data calculated from EMR data were also evaluated on the same internal validation and time-wise testing sets to ensure fair and uniform comparison across all models. Data processing and cleaning as well as model development in this study were performed using R software. RF model development was performed using python.

### Model evaluation

Multiple predictive models were built to predict surgical case duration. Different standards are usually applied to evaluate the predictive performance of the built models. The three key metrics used to evaluate model performance in this study included (1) R-square (*R*^2^), (2) mean absolute error (MAE), and (3) the percentage overage, underage and within.

*R*^2^ is the coefficient of determination; it represents the proportion of the variance for the actual case duration that is explained by predictor variables in our models.

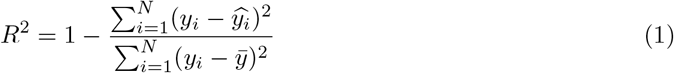

MAE measures the average of errors between the actual case durations and the predictions.

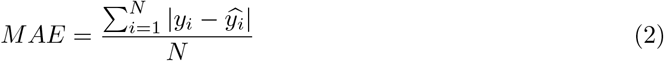

In this study, percentage within indicates the percentage of cases with absolute duration differences falling within the threshold 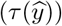 of minimum(maximum(15 %,15 min), 60 min). The mathematical expression of case duration differences that satisfy the within condition is shown as below:

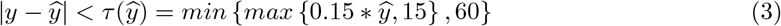

Meanwhile, percentage underage is the percentage of cases with actual case duration shorter than prediction and case duration difference was more negative than the threshold. Similarly, percentage overage is the percentage of cases with actual case duration longer than prediction and case duration difference was more positive than the threshold. The condition that defines a case as overage, within and underage is summarised as follows:

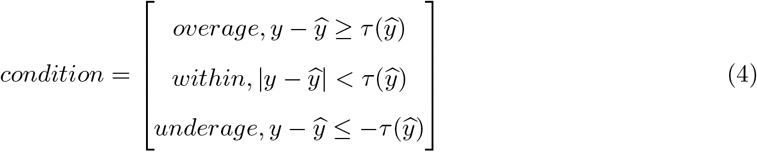

### Feature importance based on weighted feature gain

During the development of XGB model, feature importance type of ‘gain’ was applied. ‘Gain’ is the most relevant attribute to interpret the relative importance of each feature. In XGB algorithm, ‘gain’ is the average gain across all splits where feature was used and is determined by the improvement in accuracy brought by a feature to the branches it is on [30]. The calculation of ‘gain’ can be shown as below:

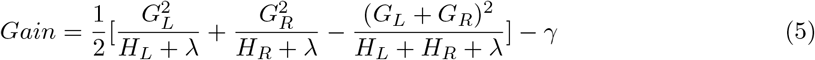

In the above equation, G is the first derivative of training loss and H is the second derivative of training loss. *λ* represents the regularization rate while *γ* represents the minimum loss reduction required to make a split. Overall, this equation consists of the score on the new left (L) leaf, the score on the new right (R) leaf, the score on the original leaf, and regularization on the additional leaf. Hence, if ‘gain’ is 0 or negative (i.e. =*< γ*), it indicates better not to add that branch. When analyzing feature importance of the model, the weighted feature gain was calculated by multiplying the percentage gain of each feature by their percentage frequency of occurrence in the model [15].

### Model interpretation with SHAP

To further interpreter the developed XGB model, we determined the Shapley additive explanations (SHAP) value by applying SHAP package in Python. SHAP can be used to explain the predicted output by computing the contribution of each feature to the prediction [31, 20]. SHAP value of a feature i, termed *φ*_*i*_, can be obtained with the following equation:

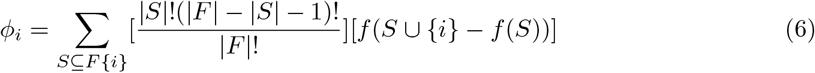

In the above equation, F is the set of all features considered for the XGB algorithm, S denotes a subset of features obtained from the set F except feature i, and *f* (*S*) is the expected output given by the set S of features. In summary, SHAP values indicate the impact of a feature on the model output. For our ML model, a large positive (negative) SHAP value of a feature implies that this feature has a large contribution in predicting a longer (shorter) surgical case duration. Meanwhile, a SHAP value of 0 implies that this feature have no or low contribution in predicting surgical case duration. SHAP values are expressed in log-odds (7) in this study.

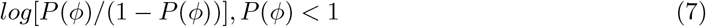

Local interaction effects between features were identified by applying SHAP interaction values in the SHAP package. While SHAP value is the attribution for each feature, SHAP interaction value is a matrix of feature attributions [32]. The interaction effects on the off-diagonal and the main effects are on the diagonal. The SHAP interaction values is defined as:

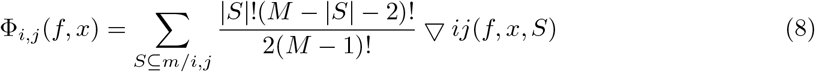

when i ≠ j and:

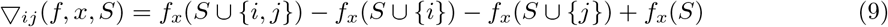

In the equation (8), m is the set of all M input features. More details regarding the computation of SHAP interaction values can be referred to Lundberg et al. (2020). The SHAP interaction value between feature i and feature j is split equally between each feature (Φ_*i,j*_(f, x) = Φ_*j,i*_(f, x)), and the total interaction effect is the sum of Φ_*i,j*_(f, x) and Φ_*j,i*_(f, x).

SHAP values of a model’s loss function decompose the model loss into the loss for each model input feature [32]. They provide information on how each feature contributes to the model loss. The definition of the sum of SHAP loss value of all features is shown as below [33]:

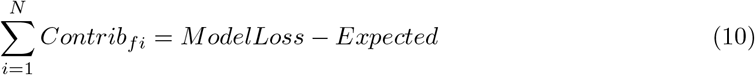

In the above equation, the calculation of expected loss is performed by first setting all the data labels to the average of all predictions, and then computes the average loss. *Contrib*_*fi*_ represents the contribution of each feature to the model loss from the expected loss.

## Data Availability

The minimum data set (March to April 2020) used in time-wise evaluation for this study is available from our web site: https://cmuhopai.azurewebsites.net/. The data set required to replicate model training and internal evaluation contains personal data and is not publicly available, in keeping with the Data Protection Policy of CMUH.

## Data availability

### Code availability

The code for data processing and model development in this study are presented on github: https://github.com/AII-CMUH/AI-surgery-scheduling.

## Acknowledgments

The authors would like to thank Min-Hsuan Lu and the department of administration and management in CMUH in the assistance of data collection.

## Competing interests

The authors declare that they have no competing interests.

## Materials & correspondence

Correspondence and requests for materials should be addressed to Jiaxin Yu.

## Supplementary Info

**Figure S1:**
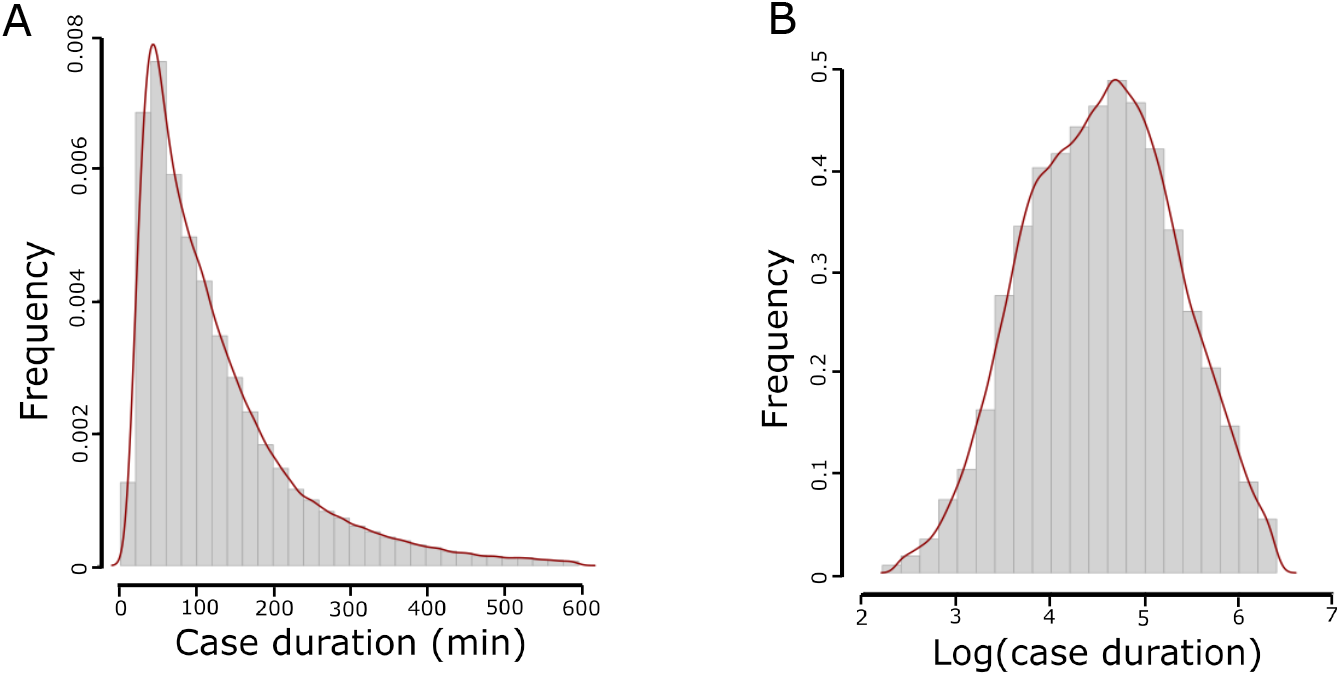
Log transformation of case duration converted the distribution of surgical case duration from (A) skewing to the right to (B) a more normal distribution.

**Figure S2:**
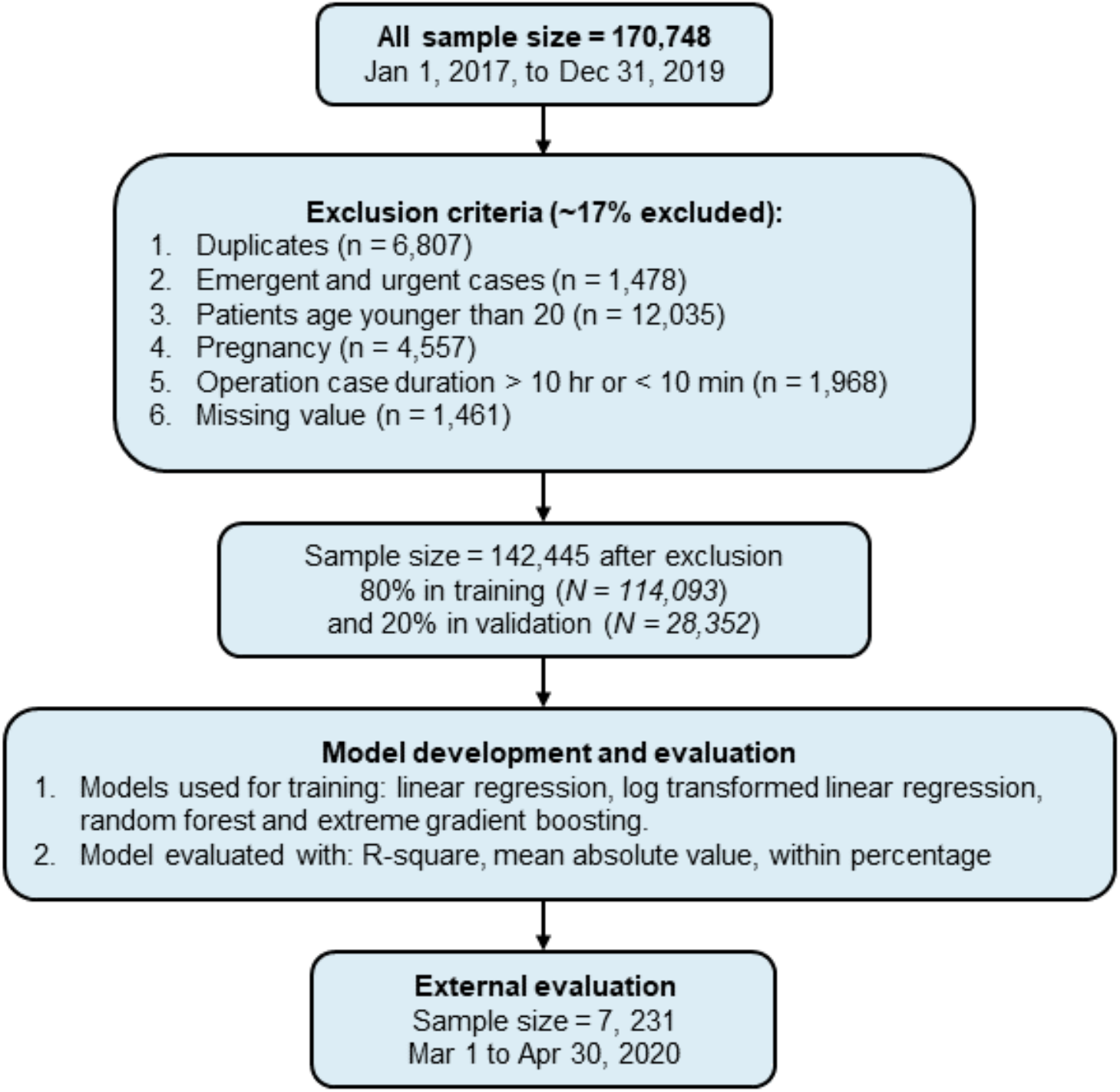
The workflow of model training for this study. The data used for model training fall within the time range of Jan 1, 2017, to Dec 31, 2019. From this data set, approximately 17 % of the cases were excluded based on the following criteria: duplicates, emergent and urgent cases, patients with age younger than 20, pregnant patients, procedure duration longer than 10 hours or less than 10 minutes and cases with missing values. The total number of cases included in the data set for model building was 142,445. This data set was then split into training (80 %) and validation (20 %) subsets for model development. Machine learning and linear regression models were developed on the training data set and validated on the validation data set using R-square and mean absolute error. The percentage of cases with duration differences falling within the threshold of minimum(maximum(15 %, 15 min),60 min) of the actual duration was also computed. Eventually, the models were further evaluated on the most recent surgical cases (from Mar 1 to Apr 30, 2020), which were not included in the original data set for model training.

**Figure S3:**
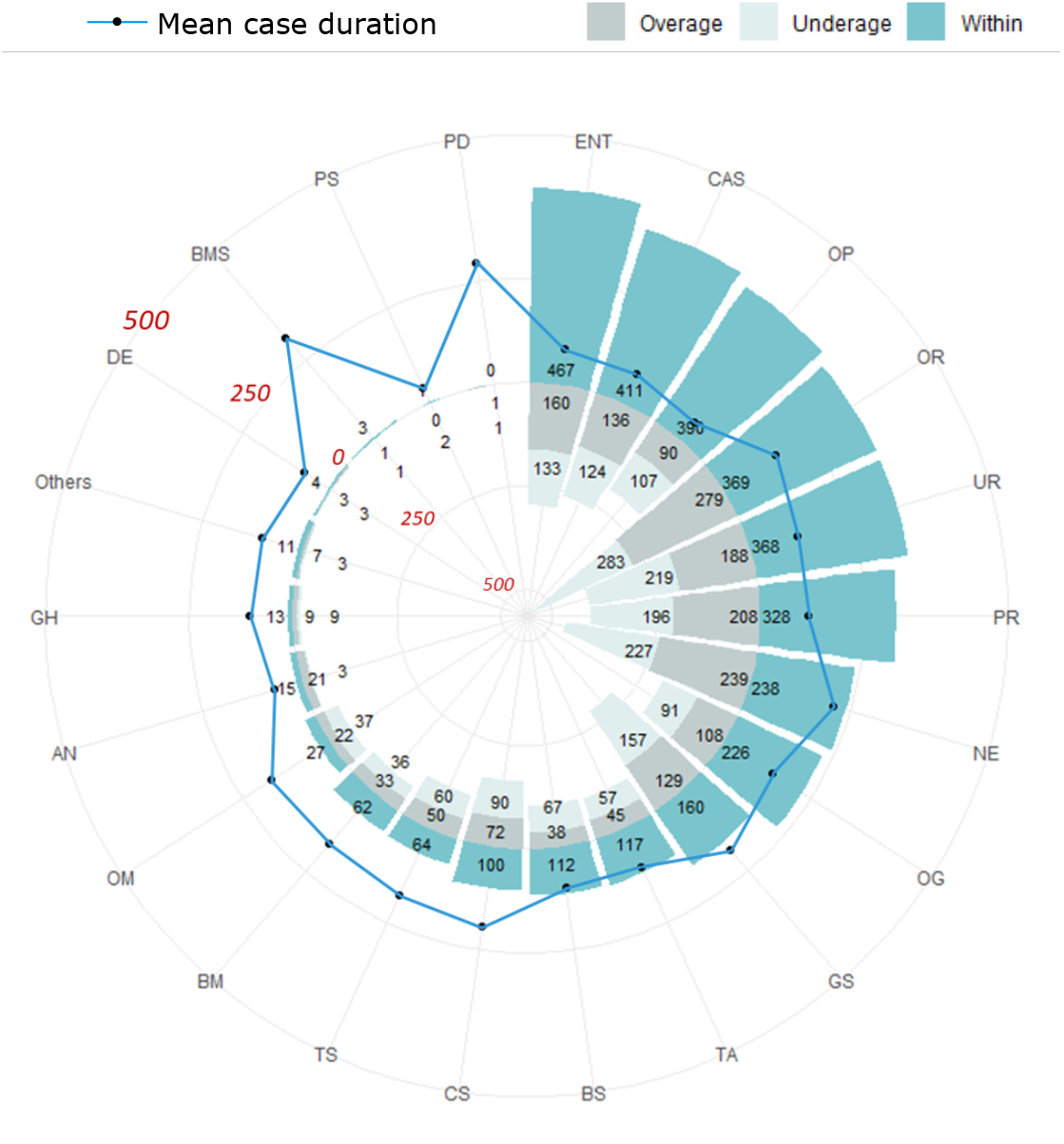
Numbers of cases that were predicted as overage, underage and within for each specialty in the time-wise testing set based on surgical case durations predicted by the extreme gradient boosting model. The black circle represents the mean case duration for each specialty. OP: ophthalmology; CAS: cardiovascular; ENT: otolaryngology, head and neck; TA: trauma and acute care; OG: obstetrics and gynecology; CS: colorectal; UR: urology; BM: body science and metabolic disorders; BS: breast surgical oncology; PR: plastic and reconstruction; GS: general; OR: orthopedics; TS: thoracic; NE: neurosurgery; OM: oral and maxillofacial; AN: anesthesiology; GH: gastroenterology and hepatology; DE: dermatology; PS: pediatric; BMS: bariatric and metabolic; PD: pediatric dentistry.

**Table S1:**
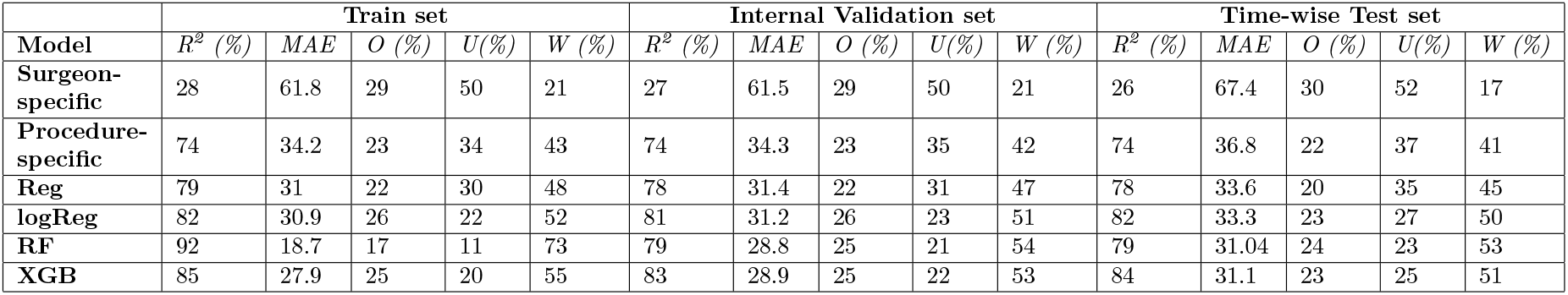
Performance of all the models in the training and internal validation and time-wise testing sets. The models that were included for comparison in this study were average models for surgeon- or procedure-specific scenarios, multivariate linear regression (Reg), log transformed multivariate linear regression (logReg), random forest (RF) and extreme gradient boosting (XGB). MAE: mean absolute error; U: underage; O: overage; W: within

|  | Train set |  |  |  |  | Internal Validation set |  |  |  |  | Time-wise Test set |  |  |  |  |
| --- | --- | --- | --- | --- | --- | --- | --- | --- | --- | --- | --- | --- | --- | --- | --- |
| Model | $R^2$ (%) | MAE | O (%) | U (%) | W (%) | $R^2$ (%) | MAE | O (%) | U (%) | W (%) | $R^2$ (%) | MAE | O (%) | U (%) | W (%) |
| Surgeon-specific | 28 | 61.8 | 29 | 50 | 21 | 27 | 61.5 | 29 | 50 | 21 | 26 | 67.4 | 30 | 52 | 17 |
| Procedure-specific | 74 | 34.2 | 23 | 34 | 43 | 74 | 34.3 | 23 | 35 | 42 | 74 | 36.8 | 22 | 37 | 41 |
| Reg | 79 | 31 | 22 | 30 | 48 | 78 | 31.4 | 22 | 31 | 47 | 78 | 33.6 | 20 | 35 | 45 |
| logReg | 82 | 30.9 | 26 | 22 | 52 | 81 | 31.2 | 26 | 23 | 51 | 82 | 33.3 | 23 | 27 | 50 |
| RF | 92 | 18.7 | 17 | 11 | 73 | 79 | 28.8 | 25 | 21 | 54 | 79 | 31.04 | 24 | 23 | 53 |
| XGB | 85 | 27.9 | 25 | 20 | 55 | 83 | 28.9 | 25 | 22 | 53 | 84 | 31.1 | 23 | 25 | 51 |

**Figure S4:**
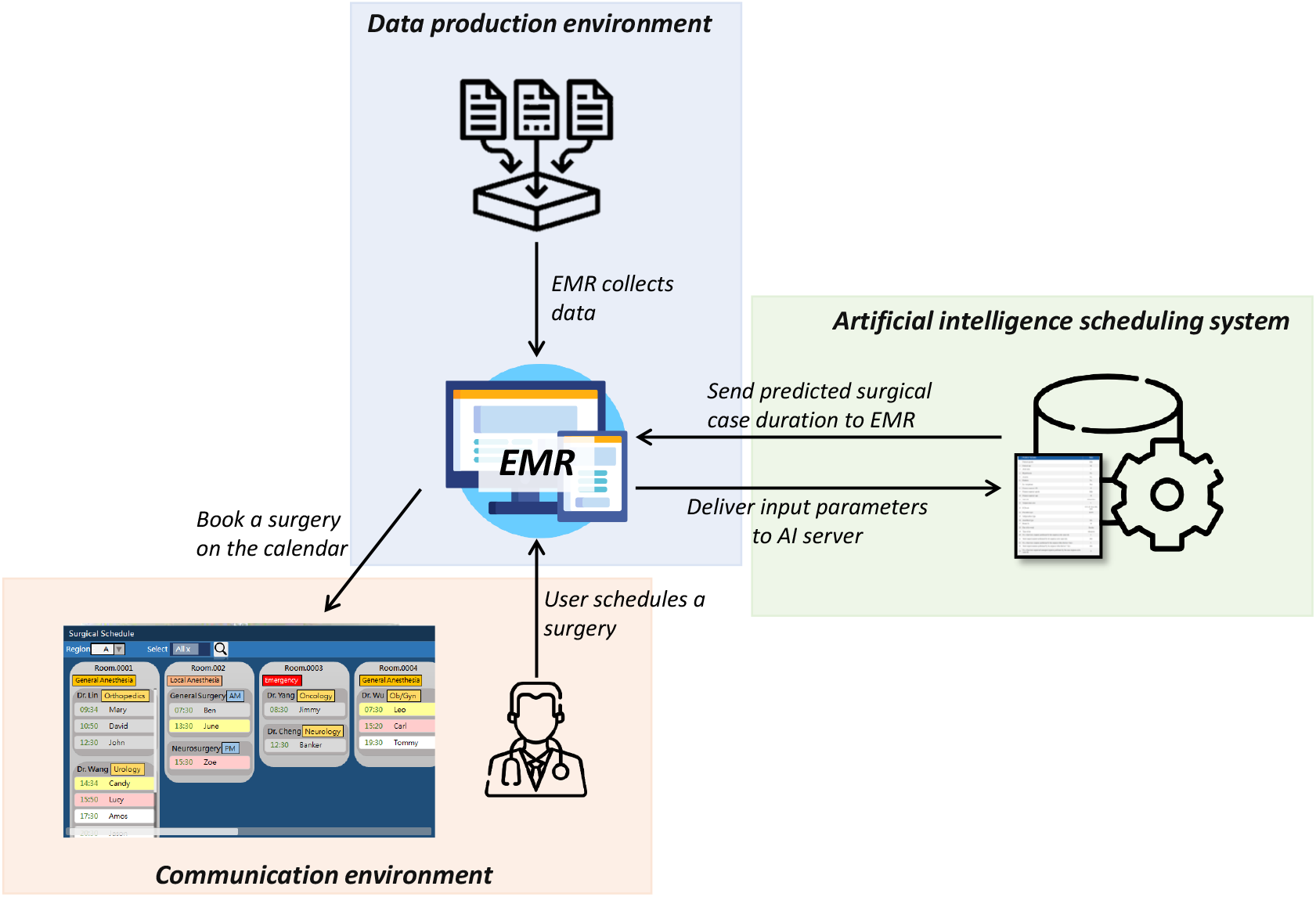
The mode of deployment of the machine learning (ML) model developed in this study. The ML model will be deployed as a stand-alone artificial intelligence (AI) server connected by the electrical medical record (EMR) system of the hospital. When a user schedules a surgery using the EMR system, the EMR delivers input parameters to the AI server, where a prediction of surgical case duration is made, and then the AI server sends the output back to the EMR as a suggestion for the user to book a surgery on the calendar.

**Table S2:** Correlation coefficient, standard error, t-value and p-value of predictor variables extracted from primary surgeons’ data. This information was obtained from the log-transformed multivariate regression (logReg) model.

| Predictor variables | Correlation coefficient | Standard error | t-value | p-value |
| --- | --- | --- | --- | --- |
| No. of previous surgeries performed by the surgeon within 24 hr | $-9.14 \times 10^{-3}$ | $7.5 \times 10^{-4}$ | -12.19 | $<2 \times 10^{-16}$ |
| Total surgical minutes performed by the surgeon within 24 hr | $6.48 \times 10^{-5}$ | $6.22 \times 10^{-6}$ | 10.41 | $<2 \times 10^{-16}$ |
| No. of previous surgeries performed by the surgeon within the last 7 days | $-1.37 \times 10^{-2}$ | $2.58 \times 10^{-4}$ | -53.35 | $<2 \times 10^{-16}$ |
| Total surgical minutes performed by the surgeon within the last 7 days | $7.71 \times 10^{-5}$ | $2.27 \times 10^{-6}$ | 33.89 | $<2 \times 10^{-16}$ |

**Table S3:** Proportion based on patient characteristics in the overall original data set and time-wise testing set.

|  | Overall (n = 142,445) | Test(n = 7,231) |
| --- | --- | --- |
| **Gender** |  |  |
| Male | 70096 (49.2%) | 3618 (50%) |
| Female | 72349 (50.8%) | 3613 (50%) |
| **Age** |  |  |
| 20-45 | 42396 (29.7%) | 2061 (28.5%) |
| 45-65 | 55134 (38.7%) | 2800 (38.7%) |
| 65-80 | 34493 (24.2%) | 1811 (25%) |
| >80 | 10422 (7.3%) | 559 (7.7%) |
| **In-/out-patient** |  |  |
| In-patient | 92780 (65.1%) | 4826 (66.7%) |
| Out-patient | 49665 (34.8%) | 2405 (33.2%) |
| **Hypertension** |  |  |
| Yes | 32398 (22.7%) | 1676 (23.1%) |
| No | 62792 (44%) | 3181 (44%) |
| Unknown | 47255 (33.1%) | 2374 (32.8%) |
| **BMI** |  |  |
| <18.5 | 33581 (23.6%) | 1945 (26.9%) |
| 18.5-22.9 | 45338 (31.8%) | 2470 (34.2%) |
| 23-26.9 | 42392 (30%) | 2246 (31.1%) |
| >= 27 | 8235 (5.8%) | 439 (6.1%) |
| Missing | 12899 (9.1%) | 131 (1.8%) |

## Notes

### Competing Interest Statement

The authors have declared no competing interest.

### Funding Statement

No external funding received.

### Author Declarations

We have obtained an institutional review board approval (CMUH109-REC1-091) from China Medical University Hospital.

### Summary of Updates

Subsection of Model interpretation under the section of Results was added to interprete the machine learning model; Fig. 2-5 were updated; authors were updated; Supplementary info was update.

